# Sleep Duration and the Prevalence of Metabolic Syndrome in adolescents and children: a Systematic Review and Meta-analysis

**DOI:** 10.1101/2022.05.11.22274958

**Authors:** Yiyang Xu, Jianian Hua, Yueping Shen

## Abstract

**Objective:** Previous studies discussing the association between sleep duration and the prevalence of metabolic syndrome have reported different results, most of which targeted at adults. We are devoted to investigating the effects of sleep time for metabolic syndrome in children conducting a meta-analysis.

**Methods:** Several cross-sectional studies were retrieved from Pubmed, Ovid, Cochrane, and Embase from inception to October 2021. Fixed-effect models and random-effect models were used to analyze the effects of sleep time on metabolic syndrome in adolescents based on the research above.

**Results:** We collected data from 10 cross-sectional studies including 15877 children. Using random-effect models, compared with normal sleep time, we found out that both relatively short (OR = 0.81, 95%CI = 0.55-1.07, I-squared = 45.6%, p = 0.075) and long (OR = 0.86, 95%CI = 0.54-1.19, I-squared = 67.8%, p = 0.003) sleep durations were not associated with high prevalence of metabolic syndrome in adolescents and children. Using fixed-effect model on short sleep duration and it turned out to be statistically significant (OR = 0.76, 95%CI = 0.59-0.93).

**Conclusions:** Long sleep duration made no significant difference in the risk of metabolic syndrome in adolescents and children, while short sleep duration seems to be a protective factor. Further studies are required to establish whether the association is causal and modifiable.

## Introduction

Metabolic syndrome (MetS) is a disease caused by a combination of many factors. It is believed as a pathological state in which the body’s protein, fat, carbohydrates, and other substances are metabolically disordered, thus being a complex of metabolic disorder syndromes and a risk factor for diabetic cardiovascular and cerebrovascular diseases. The current standards are mostly applicable to adults, for example, the National Cholesterol Education Program’s Adult Treatment Panel III (NCEP ATP-III)[1]. It defined pediatric MetS using criteria analogous to ATP III as ≥3 of the following: (1) fasting triglycerides ≥1.1 mmol/L (100 mg/dL); (2) HDL <1.3 mmol/L (50 mg/dL), except in boys aged 15 to 19 years, in whom the cutpoint was <1.2 mmol/L (45 mg/dL); (3) fasting glucose ≥6.1 mmol/L (110 mg/dL); (4) waist circumference >75th percentile for age and gender; and (5) systolic blood pressure >90th percentile for gender, age, and height[2]. According to multiple research, the prevalence of metabolic syndrome in adolescents and children is 2.0-7.6%[3-7], with increasing trends during the last decades[8]. The three most prevalent risk factors were high triglycerides, low HDL cholesterol, and obesity[6]. Another study found that adolescents with chronic kidney disease have a high prevalence of MetS[9]. Previous results have not been always consistent, so attention should be drawn to other related risk factors of metabolic syndrome in adolescents and children.

Sleep is an important part of healthy development and is necessary for physical and mental health. However, sleep duration in adolescents and children has decreased over the past century[10]. More studies have been conducted on the impact of short sleep on adolescents and children than on long sleep. These studies indicate that the short sleep duration of adolescents and children is related to high-risk behavior factors[11], type 2 diabetes[12], high HDL concentrations[13], increased abdominal adiposity, decreased, insulin sensitivity, high blood pressure[14], developing overweight and obesity[15], and long sleep duration is associated with high-risk risk of stroke in individuals with MetS[16]. A study not aimed at adolescents and children suggested long sleep is significantly associated with mortality, incident diabetes mellitus, cardiovascular disease, stroke, coronary heart disease, and obesity[17].

The association between sleep duration and the prevalence of metabolic syndrome in adults has been well established. It’s no surprise that abnormal sleep duration can cause metabolic syndrome, which is a collection of several cardiovascular risk factors. Several studies have discussed how the prevalence of metabolic syndrome is affected by sleep duration, some of which showed a U-shaped result[18, 19], which means both relatively short and long sleep duration raises the opportunity of metabolic syndrome. A recent meta-analysis found that only short sleep durations were associated with an increased risk of metabolic syndrome[20]. However, limited studies focused on adolescents and children. Existing studies have provided varied results. Studying the link between sleep time and the prevalence of metabolic syndrome in younger people is helpful to improve health guidance for underage individuals, we, therefore, performed a systematic review and meta-analysis on the association between sleep duration and the prevalence of metabolic syndrome in adolescents and children.

## Materials and Methods

We conducted this study stuck to the Preferred Reporting Items for Systematic Reviews and Meta-Analysis (PRISMA) statement[21].

Two independent researchers (YYX and JNH) separately assessed the applicability, extracted data, and assessed the quality of the included studies. Any disagreement in screening the articles was resolved through discussion between these two investigators, with consultation by a third researcher (YPS) if disagreements persisted.

### Search strategy

We performed an exhaustive search of four databases: Pubmed, Ovid, Cochrane, and Embase. The search strategy included these keywords: (”sleep duration” OR “sleep hour” OR “sleeping hour” OR “hours of sleep” OR “sleep time” OR “sleep length” OR “sleep period” OR “sleeping time”) AND (“metabolic syndrome” OR “MetS” OR “MS” OR “syndrome X” OR “cardiometabolic risk factor” OR “insulin resistance syndrome”) AND (“children” OR “child” OR “kid” OR “teen” OR “adolescent” OR “adolescence” OR “youth”). All the results were searched on Oct 4, 2021. The reference lists of retrieved articles were searched by hand.

### Inclusion criteria

All the studies included should meet the inclusion criteria: 1) focused on the association between sleep duration and the prevalence of metabolic syndrome; 2) provided the definition of metabolic syndrome and methods for measuring sleep duration as well as sufficient data to calculate odds ratio (OR) with 95% confidence interval; 3) restricted the population to adolescents and children. If more than one research reported results based on the same population, the research with the largest sample size was selected.

### Data extraction

The following information was extracted from available articles: 1) author name; 2) publication year; 3) study location (country and continent); 4) sample size; 5) sex ratio; 6) mean age with standard deviation or age range; 7) measurements of metabolic syndrome and sleep duration; 8) sleep duration category; 9) adjusted odds ratio or sufficient data for calculation.

### Quality assessment

We conducted a quality assessment of every included study using the 11-item checklist recommended by the Agency for Healthcare Research and Quality (AHRQ) because there were only cross-sectional studies included in our study after selection. Zero or one point is given for each item. AHRQ scores of <4, 4-7, and >7 were respectively considered as low, moderate, and high quality. (not given)

### Data analysis

The association between long or short sleep duration with metabolic syndrome in adolescents and children was estimated by pooled OR and 95%CI. Both random-effect models and fixed-effect models were used for calculation. Heterogeneity between studies was assessed using the Q test (P < 0.10 indicating heterogeneity was statistically significant) and I-squared statistic (I-squared > 50% indicating heterogeneity was statistically significant). A subgroup analysis was performed to explore the potential heterogeneity across cross-sectional studies after stratification by geographic region, the definition of metabolic syndrome, and measurement of sleep duration. We use the Z-test to compare the combined estimates for each subgroup.

The statistical analyses were conducted using Stata 16.0 software.

## Results

### Primary search

The initial search yielded 320 articles, among which 299 were reviewed based on the title and abstract. A total of 21 full-text articles were retrieved for final analysis. Figure 1 shows the workflow of our searching process.

**Figure 1.**
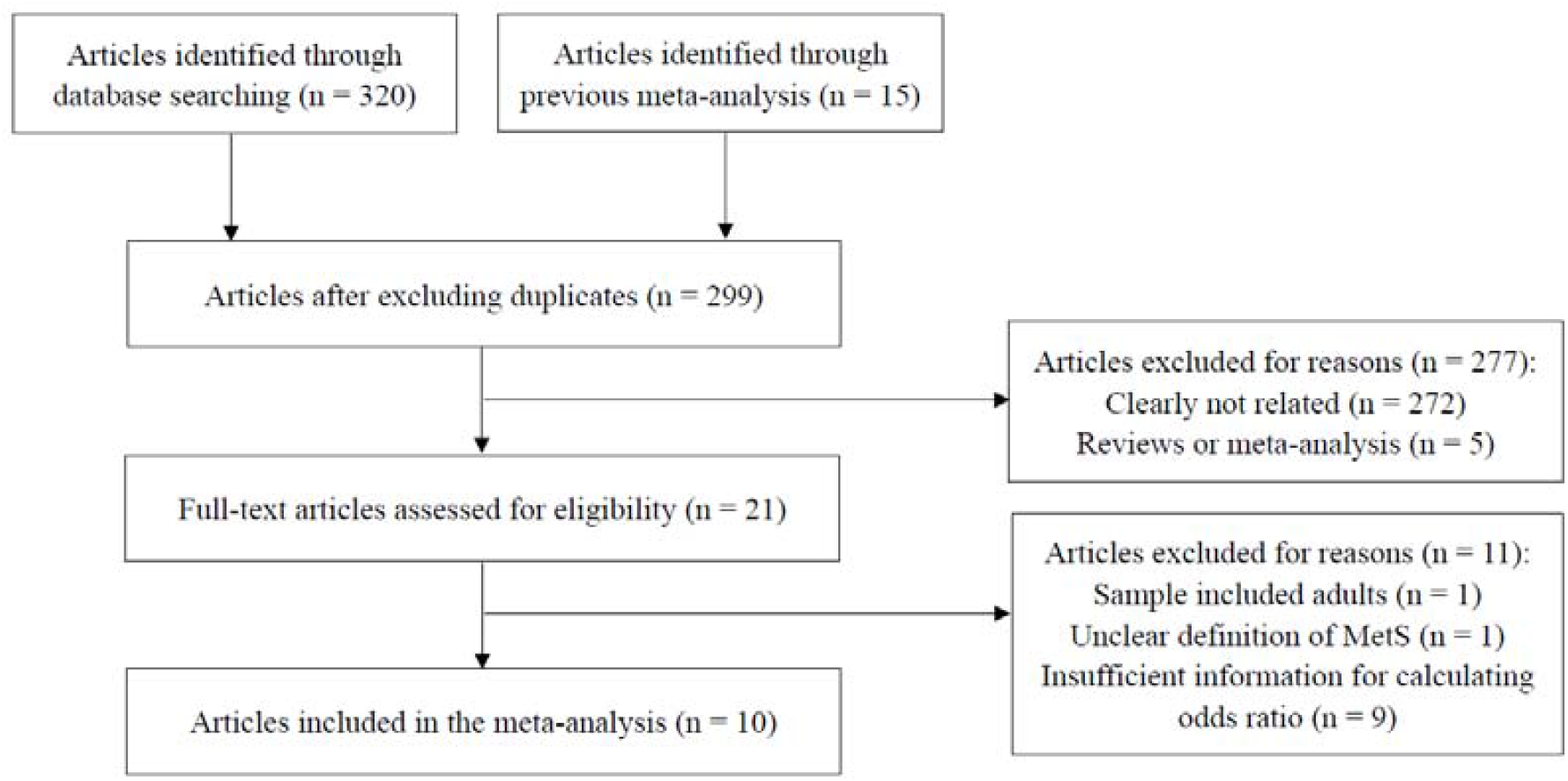
Workflow of the searching process

### Characteristics of studies

We screened out ten cross-sectional studies which discussed the relationship between sleep duration and metabolic syndrome. We didn’t find any cohort studies that met our requirements. Table 1 shows the details of each cross-sectional study.

**Table 1.**
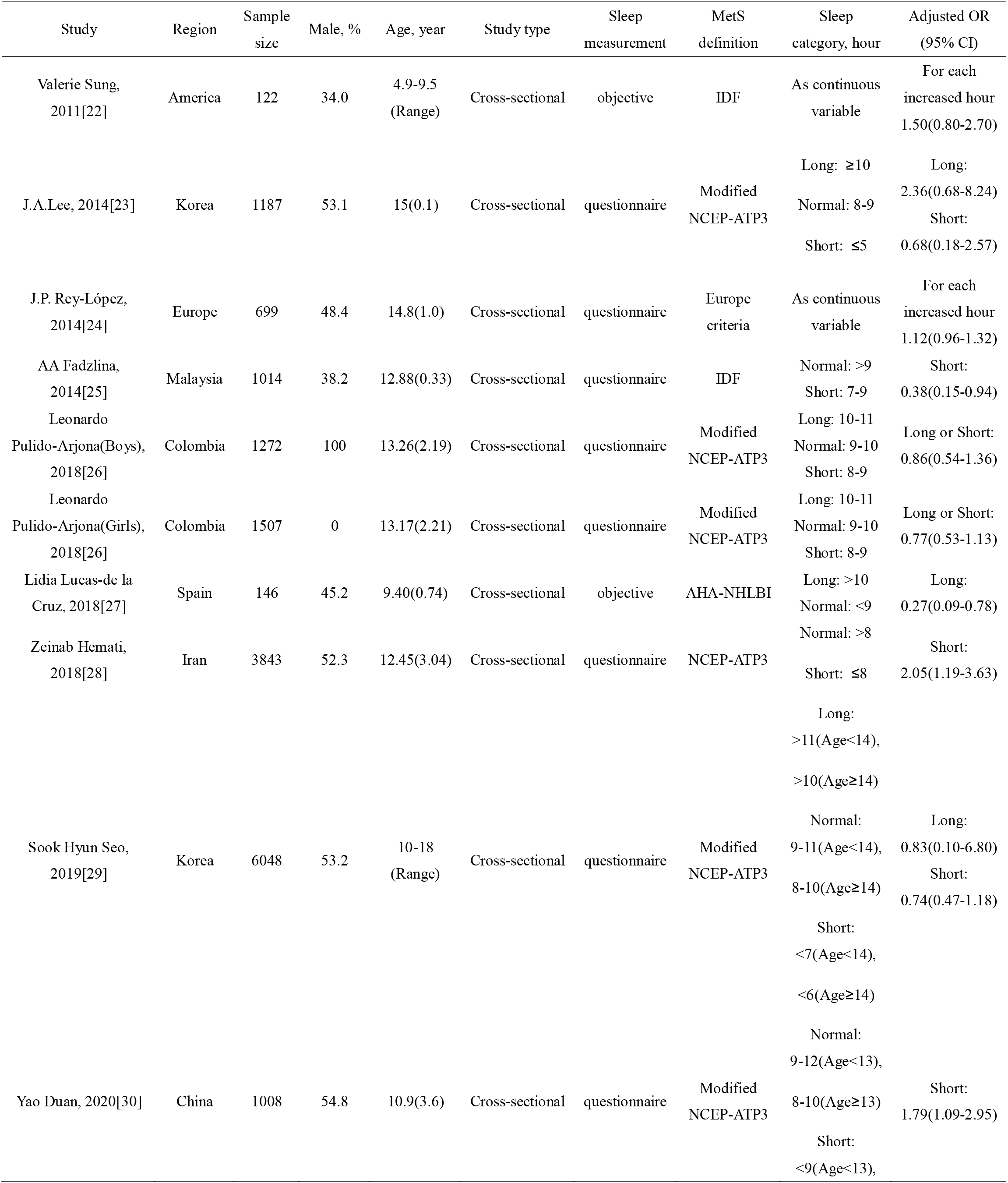
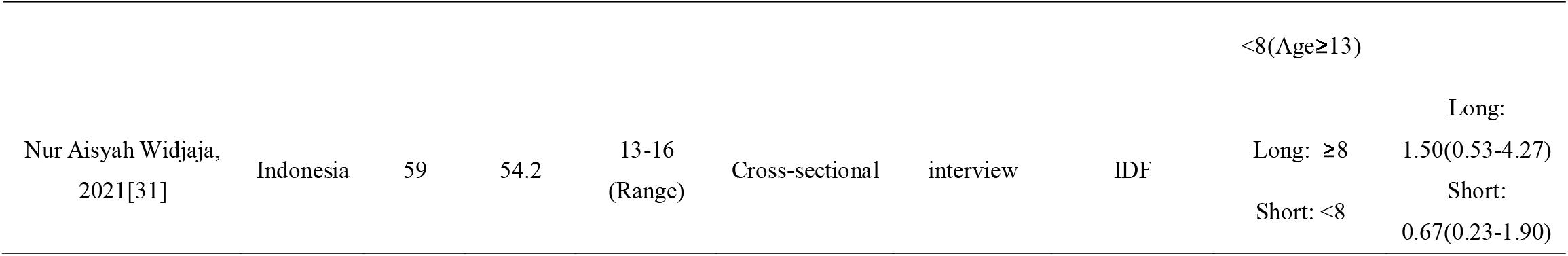
Characteristics of the studies included.

Each study involved a different number of participants, ranging from 59 to 6048, with a total of 15,877 children. These studies varied geographically, from Asia to the Americas and then to Europe, so that the races of the subjects are also not the same, thus avoiding the deviation caused by a single race. 70% of the studies used a questionnaire to measure sleep duration. Only two studies adopted the multi-night actigraphy or accelerometer as objective measurement methods[22, 27]. Moreover, it is important to note that studies differ on the definition of metabolic syndrome. NCEP-ATP III criterion is the most widely used, including its modified version. By contrast, the International Diabetes Federation(IDF) criterion[32] and the American Heart Association/National Heart, Lung, and Blood Institute(AHA-NHLBI) criterion[33] are used in fewer studies. In addition, differences exist in age range and the definition of long or short sleep duration.

### Meta-analysis

To discover the association between abnormal sleep duration and the prevalence of MetS, we used the random-effect model to conduct a meta-analysis. It turned out that compared to moderate sleep durations, both short (OR = 0.81, 95%CI = 0.55-1.07, I-squared = 45.6%, p = 0.075; Figure 2) and long (OR = 0.84, 95%CI = 0.54-1.15, I-squared = 66.1%, p = 0.004; Figure 3) sleep durations were not associated with high prevalence of metabolic syndrome in adolescents and children.

**Figure 2.**
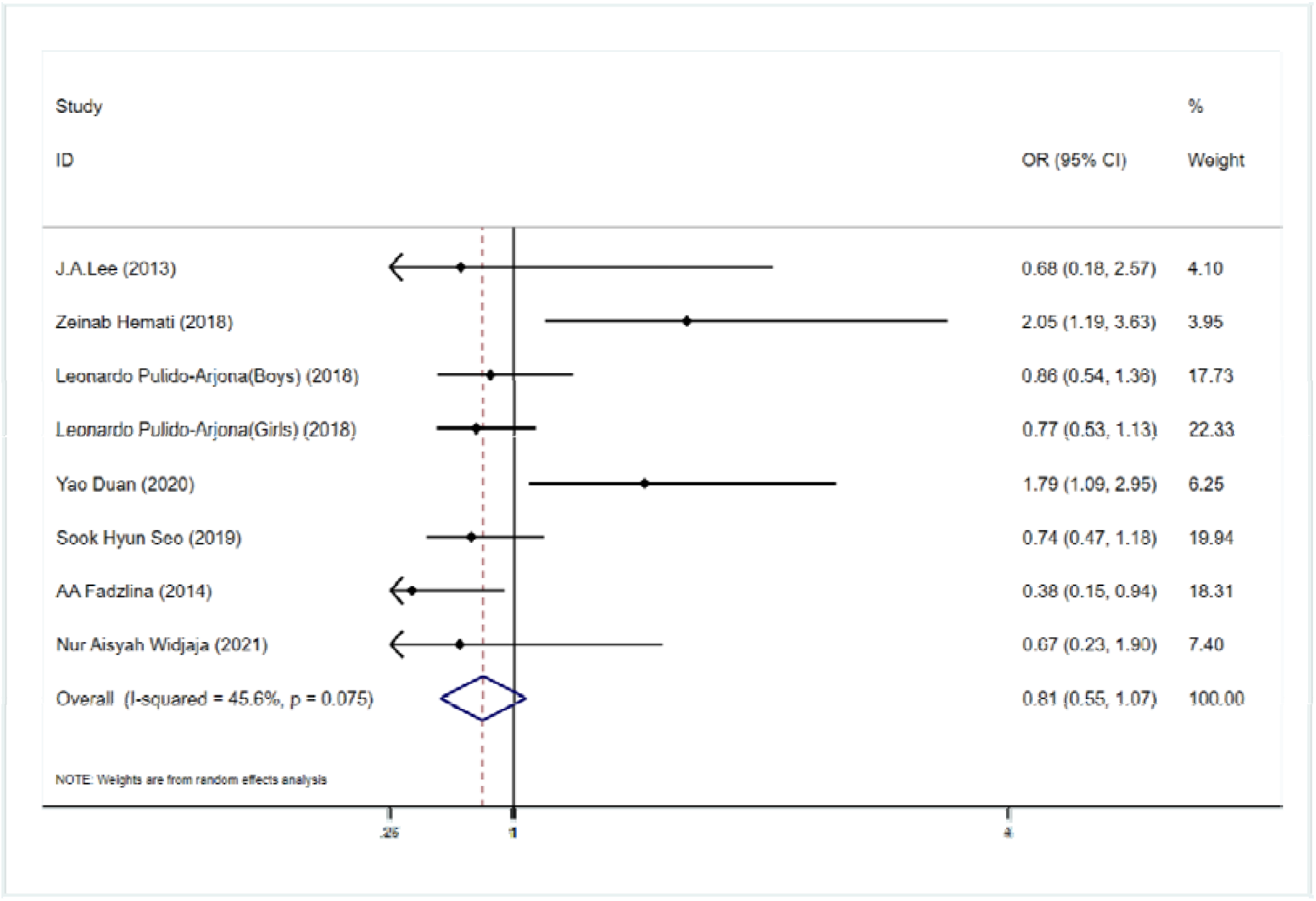
Association between short sleep duration and the prevalence of metabolic syndrome using the random-effect model

**Figure 3.**
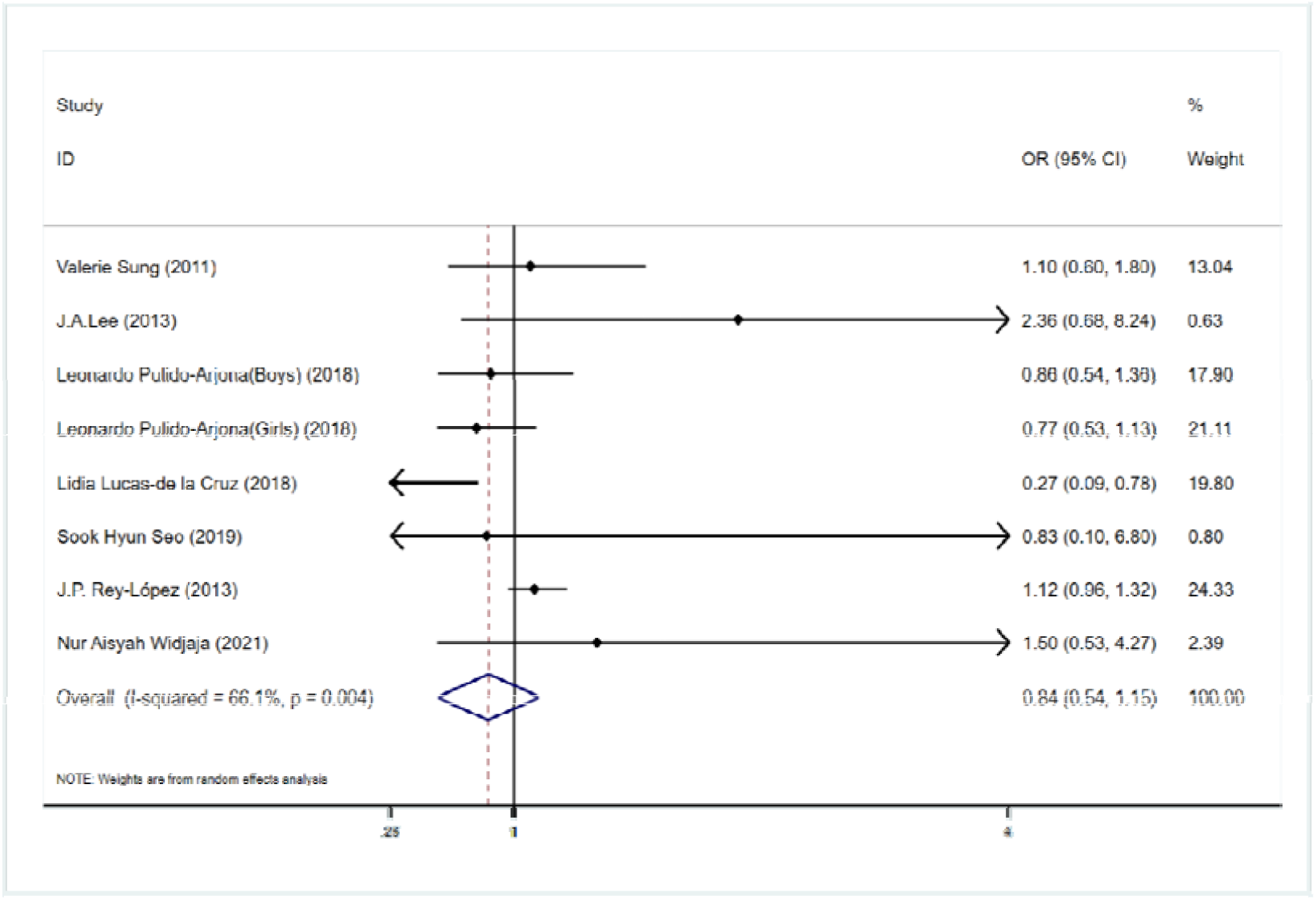
Association between long sleep duration and the prevalence of metabolic syndrome using the random-effect model

We noticed the low heterogeneity of studies that examined the effect of short sleep duration (I-squared = 45.6% < 50 %), so we conducted another meta-analysis using the fixed-effect model, and the pooled OR was 0.76 (95%CI = 0.59-0.93; Figure 4), which means for adolescents and children, relatively short sleep duration decreased the risk of metabolic syndrome.

**Figure 4.**
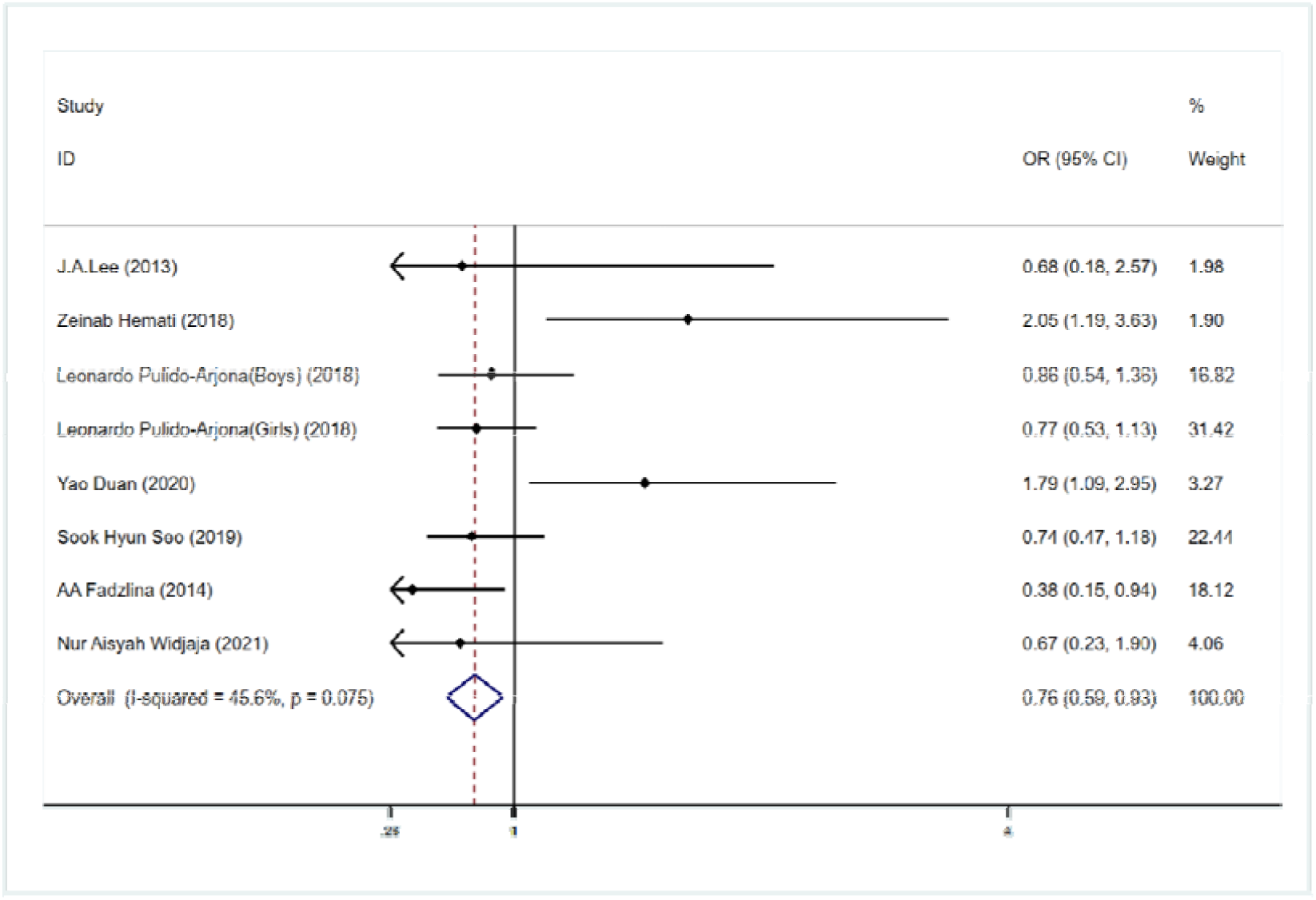
Association between short sleep duration and the prevalence of metabolic syndrome using the fixed-effect model

### Subgroup analysis

We performed some subgroup analyses based on geography, the definition of metabolic syndrome, and sleep measurement. We first divided subgroups according to the definitions of metabolic syndrome to study the effect of short sleep duration on adolescents and children. The test for subgroup differences suggested that there was a statistically significant subgroup effect (p = 0.019; Figure 5), meaning that the definition of MetS significantly modified the effect of short sleep duration on adolescents and children. Short sleep duration caused the lower risk of MetS by IDF criteria, while this effect disappeared using NECP-ATP III criteria or its modified versions; therefore the subgroup effect is qualitative. Moreover, there was no substantial unexplained heterogeneity between the studies within each of these subgroups (modified NECP-ATP III: I-squared = 12.5%; IDF: I-squared = 0%) that requires further exploration. Therefore, the validity of the short-sleep effect estimate for each subgroup was certain, as individual study results were consistent. However, both geographical location (p = 0.638) and sleep measurement (p = 0.830) showed no statistical subgroup effect.

**Figure 5.**
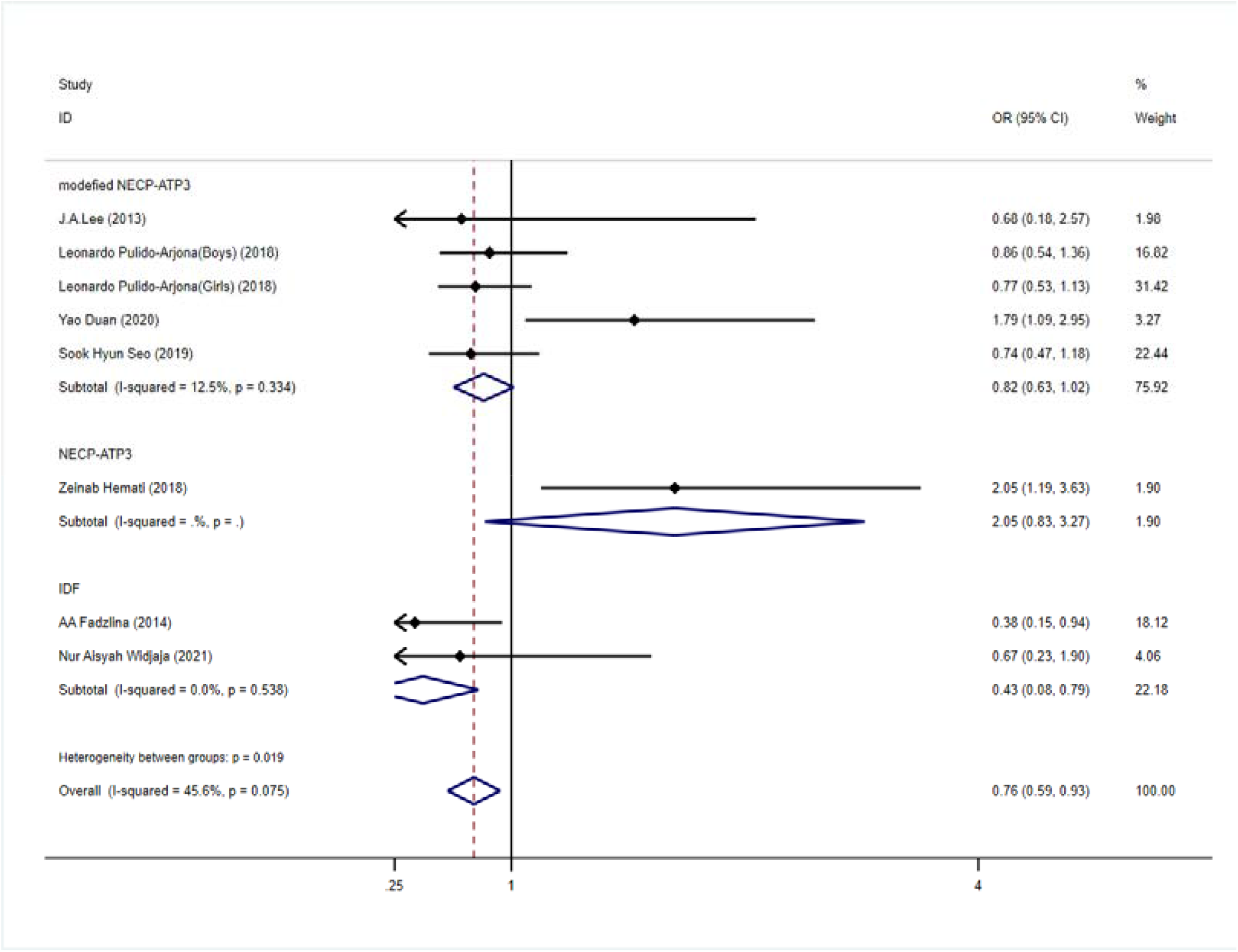
Subgroup analysis of short sleep duration by the definition of MetS

For a long sleep duration, the outcomes above displayed great heterogeneity in the meta-analysis results. Among all the subgroups stratified in different ways, two subgroups were observed to have unexplained heterogeneity (I-squared > 50%). One study they jointly included was performed by Lucas-de La Cruz, L. et al.[27]. It is suggested that the results of this study might be the source of heterogeneity in this meta-analysis.

### Sensitivity analysis and publication bias

Sensitivity analyses were conducted to explore the sources of heterogeneity among studies. According to the results (not given), none of the articles considerably shifted the effects of both short and long sleep duration on metabolic syndrome in adolescents and children. No publication bias were detected for both short sleep duration (Egger’s test: p = 0.672 and Begg’s test: p = 0.386) and long sleep duration (Egger’s test: p = 0.504 and Begg’s test: p = 1.000).

## Discussion

This study investigated the relationship between sleep duration in adolescents and children and the prevalence of metabolic syndrome. By combining the data from ten cross-sectional studies, the results suggested that while long sleep durations were not be associated with metabolic syndrome, short sleep durations were significantly associated with a lower prevalence of MetS with a pooled OR of 0.76 (95%CI = 0.59-0.93).

To our knowledge, this study represents the first meta-analysis to quantitatively investigate the association between sleep duration and the prevalence of metabolic syndrome in adolescents and children. Our findings contribute important new information to previous meta-analyses because of our new perspective, updated literature search, and the use of subgroup analysis. One meta-analysis conducted by Sun J et al. mentioned the association between sleep duration and metabolic syndrome in adolescents and children[34]. For various reasons, they did not conduct a quantitative meta-analysis of this, thus being unable to find an exact link.

By conducting a subgroup analysis, we noticed that the definition of metabolic syndrome led to modification to the effects. The OR of studies conducted in Asia was not different from studies performed on other continents. Included studies have varied definitions of short, normal, and long sleep duration. In a few studies, criteria vary by age. The results we made that are contrary to that of previous studies might be related to it. Objective measures of sleep duration are considered more reliable than subjective measures. We did not observe differences between sleep durations recorded by questionnaires or objective measures. This association was not the same as the “U-shaped” association between sleep duration and MetS reported in many articles. A reasonable explanation is the small number of extreme data, that is, the small number of children with very short or very long sleep duration, resulting in the odds ratio at both ends of the possible U-shaped association not exceeding 1, which in turn results in the failure of short or long sleep to show the role of risk factors, and even manifested as protective factors.

Several biological mechanisms linked sleep duration to metabolic syndrome. Short sleep duration may contribute to the endocrine changes described below by affecting carbohydrate metabolism, sympathetic nerve activity, and the hypothalamic-pituitary-adrenal axis. Reduced glucose tolerance and insulin sensitivity increase glucose levels; increased ghrelin, decreased leptin, and increased appetite are associated with increased waist circumference; and increased cortisol concentrations are associated with increased blood pressure[35, 36]. People with short sleep duration tend to have elevated levels of high-sensitivity C-reactive protein, which have been associated with cardiovascular events[37, 38]. Both short and long sleep duration show bidirectional association with circadian rhythm, a risk factor for metabolic disturbances[39, 40].

Previous studies supported the association between sleep duration and high blood pressure[41], obesity[42-44], and psychological diseases[45, 46] in adolescents and children. These results indicate that shorter sleep duration can be detrimental to the health status of adolescents in many ways. This is highly possible to cause various metabolic disorders, leading to metabolic syndrome. However, our findings are completely contrary to previous mainstream scientific studies. Future research should explore the exact biological mechanism in depth or explain it at the genetic level.

The strengths of this meta-analysis and systematic review include rigorous use of the PRISMA statement. A detailed search strategy was used across multiple databases with broad date ranges and stringent inclusion criteria were applied during the study selection process. The included studies were performed in four different continents, avoiding racial differences.

Our study is subject to the following limitations. First, most included studies measured sleep time using subjective questionnaires rather than objective actigraphy, increasing the systematic error of the studies. Second, the criteria used to diagnose metabolic syndrome are not consistent. Modified NCEP-ATP3 is the most widely used, but its definition of metabolic syndrome is different from IDF and AHA-NHLBI standards. The existing standards are all geared towards adults, so a unified standard is required to be promulgated specifically for adolescents and children. Third, we only focus on the relationship between sleep duration and MetS instead of obstructive sleep apnea and difficulty in initiating and maintaining sleep because there is very little research on this. Further meta-analysis ought to clarify these associations. Fourth, all the included studies were cross-sectional studies because no longitudinal studies can be searched so far, which means that the exact causal relationship cannot be determined. Finally, as there were fewer than 10 studies in each meta-analysis, funnel plot analyses were not conducted because of the low power of this test when the number of included studies is small.

## Conclusion

Long sleep duration has no significant effect on the risk of metabolic syndrome in adolescents and children, while short sleep duration appears to be a protective factor. Further research is needed to establish the stability of the association.

## Data Availability

All data produced in the present work are contained in the manuscript

## Funding

## Notes

### Competing Interest Statement

The authors have declared no competing interest.

### Funding Statement

This study did not receive any funding

